# Factors Associated with Serious Psychological Distress during the COVID-19 Pandemic in Japan

**DOI:** 10.1101/2021.02.27.21252458

**Authors:** Takashi Yoshioka, Ryo Okubo, Takahiro Tabuchi, Satomi Odani, Tomohiro Shinozaki, Yusuke Tsugawa

**Affiliations:** Center for Innovative Research for Communities and Clinical Excellence (CiRC^2^LE), Fukushima Medical University, Fukushima, Japan; Department of Clinical Epidemiology, Translational Medical Center, National Center of Neurology and Psychiatry, Tokyo, Japan; Cancer Control Center, Osaka International Cancer Institute, Osaka, Japan; Department of Information and Computer Technology, Faculty of Engineering, Tokyo University of Science, Tokyo, Japan; Division of General Internal Medicine and Health Services Research, David Geffen School of Medicine at UCLA, Los Angeles, CA, USA; Department of Health Policy and Management, UCLA Fielding School of Public Health, Los Angeles, CA, USA

## Abstract

**Importance:** The coronavirus disease 2019 (COVID-19) pandemic may have a negative impact on mental health of the population, leading to higher suicide rates, in many countries. However, little is known about risk factors associated with worsened mental health during the COVID-19 pandemic.

**Objective:** To investigate the factors associated with serious psychological distress (SPD) during the COVID-19 pandemic in Japan.

**Design, Setting, and Participants:** A cross-sectional study using a large-scale internet survey conducted between August 25 and September 30, 2020, in Japan.

**Exposures:** Demographics (age, gender, marital status, family composition, and caregiving burden), socio-economic status (income level, employment type, educational attainment), the experience of domestic violence (DV), the state of emergency, fear of COVID-19, and stigma related to COVID-19.

**Main Outcomes and Measures:** Prevalence of SPD, defined as Kessler 6 scale score ≥13.

**Results:** Among 25,482 individuals included in this study, 2,556 (10%) met the criteria of SPD. Overall, women (adjusted odds ratio [aOR] 1.59; 95%CI, 1.17–2.16; P=0.003), ages 15–29 (aOR compared with ages 45–59, 2.35; 95%CI, 1.64–3.38; P<0.001), low income (aOR compared with intermediate income, 1.70; 95%CI, 1.16–2.49; P=0.007), providing caregiving to family members (aOR, 5.48; 95%CI, 3.51–8.56; P<0.001), experiencing DV (aOR, 5.72; 95%CI, 3.81–8.59; P<0.001), and fear of COVID-19 (aOR, 1.96; 95%CI, 1.55–2.48; P<0.001) were associated with a higher rate of SPD. Among women aged 15–29 years, who experienced the highest rate of SPD, caregiving, DV, fear of COVID-19, and COVID-19-related stigma were associated with a higher rate of SPD; whereas economic situation (income level and employment type) and social isolation (marital status) were not associated with the prevalence of SPD.

**Conclusions and Relevance:** Economic situation, caregiving burden, DV, and fear of COVID-19 were independently associated with SPD during the COVID-19 pandemic. Among young women—who have a higher risk of suicide during the COVID-19 pandemic in Japan—similar factors, except economic situation, were associated with a higher rate of SPD. Targeted interventions based on age and gender may be more effective in mitigating the negative impact of the COVID-19 pandemic on the population’s mental health.

## INTRODUCTION

The coronavirus disease 2019 (COVID-19) has infected more than 110 million people, contributed to over 2.4 million deaths globally,^1^ and has impacted many aspects of our lives. The COVID-19 pandemic has put a significant burden on our healthcare systems, and many countries have been struggling with an economic downturn due to reduced economic activities or lockdowns, leading to widened social inequalities.^2^ Reduced social interactions due to social distancing and isolation, as well as economic downturns due to the pandemic, have the potential to negatively impact mental health conditions leading to psychological distress and increased risk for psychiatric disorders.^3,4^ Some countries have been experiencing an increased suicide rate during the COVID-19 pandemic. For example, in Japan, the number of people who died of suicide increased from 20,169 in 2019 to 20,919 in 2020.^5^ It is important to note that the suicide rate has increased dramatically among young women during the COVID-19 pandemic,^6^ and some have speculated that the economic downturn combined with a relatively unstable employment status often experienced among this population may be underlying reasons for increased suicides among this population. Although there are people who commit suicide without experiencing psychological distress, it is clearly one of the major risk factors for suicide.^7^ Therefore, identifying risk factors associated with psychological distress is critically important for policymakers to design interventions that can effectively mitigate deteriorating mental health conditions.

Research has found that those who are vulnerable to financial stressors (e.g., low-income levels or unemployed) are associated with psychological distress during the COVID-19 pandemic.^8–10^ Notably, women and the young are more likely to experience psychological distress,^10^ although the risk of severe disease and death due to COVID-19 is higher in men and older adults.^11,12^ This may be attributable to the difference in the prevalence of psychological distress. For example, depression and anxiety are generally more prevalent among women than men.^13^ However, specific risk factors associated with psychological distress in women—except the difference in the prevalence of depression and anxiety—remain unknown, making it difficult for policymakers to design interventions that can effectively mitigate deteriorating mental health conditions in the population. Moreover, to our knowledge, no study to date has investigated the factors associated with psychological distress among young women who are experiencing a higher risk of suicide during the COVID-19 pandemic.

In this context, using data from a large-scale internet survey conducted during the COVID-19 pandemic (between August 25 and September 30, 2020) in Japan, we sought to answer two key questions: What are the factors associated with a higher prevalence of serious psychological distress (SPD) during the COVID-19 pandemic? What are the risk factors of SPD among young women who are exhibiting a dramatic increase in suicides during the COVID-19 pandemic in Japan?

## METHODS

### Data

We analyzed data from the *Japan “COVID-19 and Society” Internet Survey (JACSIS)*. The JACSIS is a large-scale, internet-based, self-reported questionnaire survey via a survey panel provided by a major internet survey agency in Japan (Rakuten Insight, Inc., Tokyo, Japan).^14^ The total number of individuals included in the survey panel was approximately 2.2 million and comprises individuals from diverse socio-economic backgrounds—such as educational level, household income and number of household members, and marital status—to be nationally representative.^15^ This study reached out to 224,389 participants using a stratified sampling approach by gender, age, and each prefecture from the panel. The study enrollment continued until it achieved the target numbers of respondents whose age, gender, and prefectures had been *a priori* set (based on the distribution of the general Japanese population in 2019; and 28,000 respondents). The survey was conducted between August 25 and September 30, 2020. The overall response rate accounted for 12.5% (28,000/224,389). This study excluded respondents whose answers were inconsistent (we included specific items in the survey to identify inconsistent responses).

### Exposure variables

Our exposure variables of interest were respondents’ demographics and socio-economic status (SES),^16^ the experience of domestic violence (DV),^17^ the state of emergency (SOE) in response to COVID-19 (**Supplementary Method**),^18^ fear of COVID-19,^19^ and stigma related to COVID-19.^20^ The demographics included age groups (15–29, 30–44, 45–59, 60–79 years), gender,^16^ marital status (unmarried, married, and widowed/separated), having children, and caregiving to an elderly/disabled family member.^21^ The SES included educational attainment (high-school-educated or lower, and college-educated or higher),^22^ income level (categorized by the tertiles of household equivalent income [low, <2.5 million JPY; intermediate, 2.5–4.3 million JPY; high, 4.3< million JPY; unknown/declined to answer]),^23^ employment type (employer, self-employed, regular employee, non-regular employee, and unemployed).^24^ The experiences of DV (including any of physical, sexual, and financial violence) and COVID-19-related stigma were defined using the specific survey questions (**Supplementary Method**). Fear of COVID-19 was categorized by the median of the fear of COVID-19 scale (FCV-19S),^25^ which is validated in Japanese.^26^

### Adjustment variables

Adjustment variables were health-related status as follows: smoking status (never, ever, and current smokers), alcohol use (never, ever, and current users), and comorbidities. The comorbidities included those at risk of severe COVID-19, which may influence mental health during the COVID-19 pandemic (hypertension, diabetes, asthma/chronic obstructive pulmonary disease [COPD], cardiovascular disease, stroke, cancer, and psychiatric disorders).^27,28^

### Outcome variable

The outcome variable is SPD, defined as Kessler 6 scale (K-6) score ≥13.^29,30^ K-6 was validated in Japanese.^31^ SPD is a relevant outcome for policy because it is strongly associated with mental health services.^32^

### Statistical analysis

First, for each gender, we described the exposure variables (demographics, SES, experience of DV, SOE in relation to COVID-19, fear of and stigma related to COVID-19) and the adjustment variables (smoking status, alcohol use, and comorbidities) with number and proportion (%) of each category.

Second, we conducted multivariable logistic regression analyses to determine the potential risk factors of SPD among exposure variables. To characterize the risk factors for young women in addition to the general public, we also conducted multivariable logistic regression analysis focusing on women aged 15–29 years whom prior reports have shown a dramatic increase in the suicide rate in Japan during the COVID-19 pandemic ^6^ Regarding this analysis, some comorbidities (hypertension, diabetes, COPD, cardiovascular disease, stroke, cancer) were uncommon among young women, and therefore, were not included as adjustment variables.

To account for the differences between the sociodemographic status in actual respondents from the survey panel and the Japanese general public, we used inverse sampling-probability weighting for all analyses.^33,34^ The sampling weights using propensity scores were calculated by fitting a logistic regression model using sociodemographic and health-related characteristics to adjust for the difference in respondents between the current internet survey and the 2016 Comprehensive Survey of Living Conditions, which is a nationally representative survey in Japan.^33^

Statistical significance was set at P<0.05. The data were analyzed using STATA version 16.1 (Stata Corp., College Station, TX, USA). We obtained web-based informed consent from all respondents. The Institutional Review Board of Osaka International Cancer Institute approved this study (No. 20084).

## RESULTS

### Characteristics of respondents

After excluding 2,518 respondents who met the exclusion criteria, a total of 25,482 respondents (91.0% of the overall survey respondents) were included in our study. Of the overall weighted respondents, 50.3% were women; 27.2% were ages 45–59; 51.2% were college-educated or higher; 28.8% were low-income level; 63.2% were currently married; and 30.9% were regular employees (**Table 1**). The characteristics of overall unweighted respondents and those aged 15–29 years were shown in **Supplementary Table 1** and **Supplementary Table 2**, respectively.

### Factors associated with SPD in overall respondents

Of the overall weighted respondents, SPD was prevalent in 10.0% of the individuals (which amounts to 2,556 cases among 25,482 weighted total). After adjusting for smoking status, alcohol use, and comorbidities, women (adjusted odds ratio [aOR], 1.59; 95% confidence interval [CI], 1.17–2.16; P=0.003); aged 15–29 (aOR, 2.35 compared with ages 45–59 years; 95%CI, 1.64–3.38; P<0.001) and 30–44 (aOR, 1.67; 95%CI, 1.35–2.08; P<0.001); low-(aOR, 1.70 compared with intermediate-income; 95%CI, 1.16–2.49; P=0.007) and high-income level (aOR, 1.74; 95%CI, 1.25–2.42; P=0.001); self-employed (aOR, 2.11 compared with regular employee; 95%CI, 1.21–3.68; P=0.008); caregiving to an elderly/disabled family member (aOR, 5.48; 95%CI, 3.51–8.56; P<0.001); experiencing DV (aOR, 5.72; 95%CI, 3.81–8.59; P<0.001); and fear of COVID-19 (aOR, 1.96; 95%CI, 1.55–2.48; P<0.001) were independently associated with higher odds of experiencing SPD (**Table 2**)

**Table 1.**
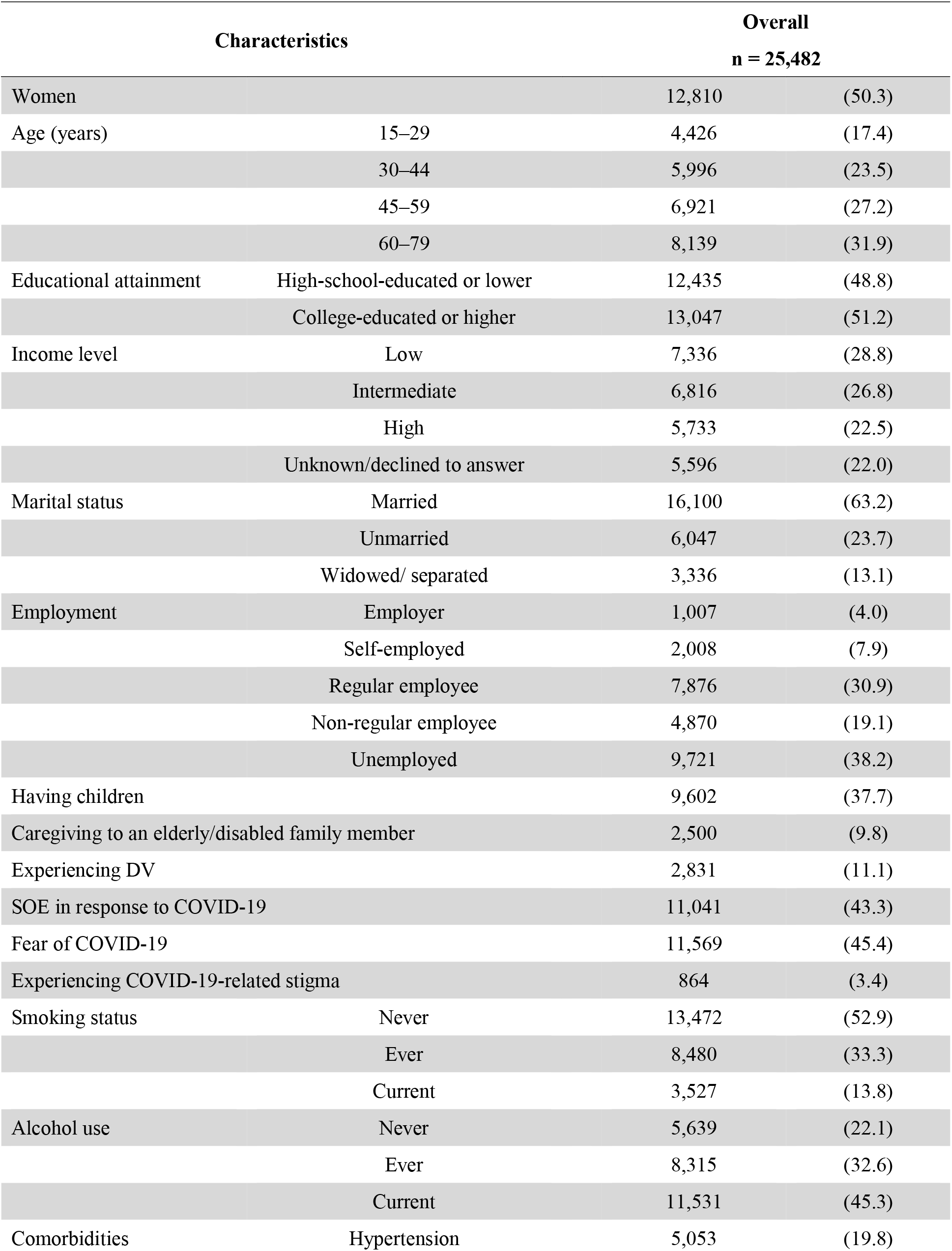
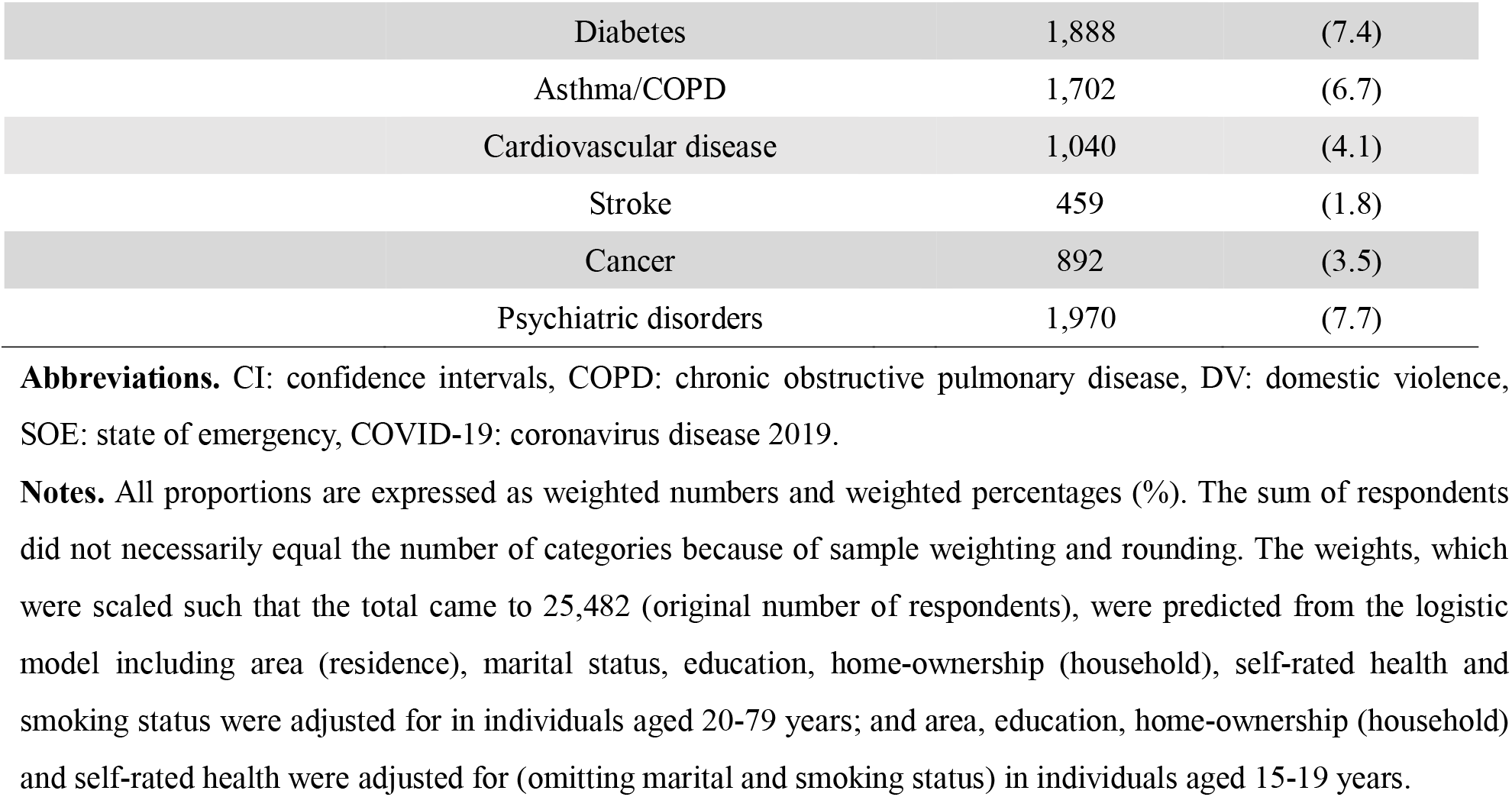
Baseline characteristics of weighted respondents.

| Characteristics |  | Overall<br>n = 25,482 |  |
| --- | --- | --- | --- |
| Women |  | 12,810 | (50.3) |
| Age (years) | 15–29 | 4,426 | (17.4) |
|  | 30–44 | 5,996 | (23.5) |
|  | 45–59 | 6,921 | (27.2) |
|  | 60–79 | 8,139 | (31.9) |
| Educational attainment | High-school-educated or lower | 12,435 | (48.8) |
|  | College-educated or higher | 13,047 | (51.2) |
| Income level | Low | 7,336 | (28.8) |
|  | Intermediate | 6,816 | (26.8) |
|  | High | 5,733 | (22.5) |
|  | Unknown/declined to answer | 5,596 | (22.0) |
| Marital status | Married | 16,100 | (63.2) |
|  | Unmarried | 6,047 | (23.7) |
|  | Widowed/ separated | 3,336 | (13.1) |
| Employment | Employer | 1,007 | (4.0) |
|  | Self-employed | 2,008 | (7.9) |
|  | Regular employee | 7,876 | (30.9) |
|  | Non-regular employee | 4,870 | (19.1) |
|  | Unemployed | 9,721 | (38.2) |
| Having children |  | 9,602 | (37.7) |
| Caregiving to an elderly/disabled family member |  | 2,500 | (9.8) |
| Experiencing DV |  | 2,831 | (11.1) |
| SOE in response to COVID-19 |  | 11,041 | (43.3) |
| Fear of COVID-19 |  | 11,569 | (45.4) |
| Experiencing COVID-19-related stigma |  | 864 | (3.4) |
| Smoking status | Never | 13,472 | (52.9) |
|  | Ever | 8,480 | (33.3) |
|  | Current | 3,527 | (13.8) |
| Alcohol use | Never | 5,639 | (22.1) |
|  | Ever | 8,315 | (32.6) |
|  | Current | 11,531 | (45.3) |
| Comorbidities | Hypertension | 5,053 | (19.8) |

|  |  |  |
| --- | --- | --- |
| Diabetes | 1,888 | (7.4) |
| Asthma/COPD | 1,702 | (6.7) |
| Cardiovascular disease | 1,040 | (4.1) |
| Stroke | 459 | (1.8) |
| Cancer | 892 | (3.5) |
| Psychiatric disorders | 1,970 | (7.7) |

**Abbreviations.** CI: confidence intervals, COPD: chronic obstructive pulmonary disease, DV: domestic violence, SOE: state of emergency, COVID-19: coronavirus disease 2019.
**Notes.** All proportions are expressed as weighted numbers and weighted percentages (%). The sum of respondents did not necessarily equal the number of categories because of sample weighting and rounding. The weights, which were scaled such that the total came to 25,482 (original number of respondents), were predicted from the logistic model including area (residence), marital status, education, home-ownership (household), self-rated health and smoking status were adjusted for in individuals aged 20-79 years; and area, education, home-ownership (household) and self-rated health were adjusted for (omitting marital and smoking status) in individuals aged 15-19 years.

**Table 2.** Factors associated with serious psychological distress among the general population.

| Characteristics |  | aOR | (95% CI) | P value |
| --- | --- | --- | --- | --- |
| Women |  | 1.59 | (1.17–2.16) | 0.003 |
| Age (years) | 15–29 | 2.35 | (1.64–3.38) | <0.001 |
|  | 30–44 | 1.67 | (1.35–2.08) | <0.001 |
|  | 45–59 |  | Ref |  |
|  | 60–79 | 0.50 | (0.3–0.81) | 0.005 |
| Educational attainment | College-educated or higher | 1.11 | (0.86–1.43) | 0.44 |
| Income level | Low | 1.70 | (1.16–2.49) | 0.007 |
|  | Intermediate |  | Ref |  |
|  | High | 1.74 | (1.25–2.42) | 0.001 |
|  | Unknown/declined to answer | 1.19 | (0.86–1.66) | 0.29 |
| Marital status | Married |  | Ref |  |
|  | Unmarried | 1.26 | (0.88–1.81) | 0.20 |
|  | Widowed/separated | 1.45 | (0.88–2.37) | 0.14 |
| Employment | Employer | 1.11 | (0.35–3.49) | 0.86 |
|  | Self-employed | 2.11 | (1.21–3.68) | 0.008 |
|  | Regular employee |  | Ref |  |
|  | Non-regular employee | 0.92 | (0.68–1.25) | 0.59 |
|  | Unemployed | 0.75 | (0.51–1.1) | 0.14 |
| Having children |  | 0.97 | (0.7–1.35) | 0.86 |
| Caregiving to an elderly/disabled family member |  | 5.48 | (3.51–8.56) | <0.001 |
| Experiencing DV |  | 5.72 | (3.81–8.59) | <0.001 |
| SOE in response to COVID-19 |  | 1.23 | (0.98–1.56) | 0.08 |
| Fear of COVID-19 |  | 1.96 | (1.55–2.48) | <0.001 |
| Experiencing COVID-19-related stigma |  | 1.22 | (0.69–2.13) | 0.50 |

**Abbreviations:** aOR: adjusted odds ratio, CI: confidence intervals, DV: domestic violence, SOE: state of emergency, COVID-19: coronavirus disease 2019.
**Notes:** Adjusted for smoking status, alcohol use, and comorbidities (hypertension, diabetes, chronic obstructive pulmonary disease or asthma, cardiovascular disease, stroke, cancer, and psychiatric disorder). All CIs and *P* values were based on the robust variance estimator to account for the inverse probability weighting.

### Analysis focusing on women aged 15-29 years

Of 2,295 weighted women aged 15–29 years, SPD was prevalent in 15.0% individuals (which amounts to 344 cases among 2,295 weighted samples). After adjustment for potential confounders, caregiving to an elderly/disabled family member (aOR, 4.05; 95%CI, 1.69–9.75; P=0.002), experiencing DV (aOR, 3.44; 95%CI, 1.94–6.11; P<0.001), fear of COVID-19 (aOR, 1.94; 95%CI, 1.30–2.89; P=0.001), and experiencing COVID-19-related stigma (aOR, 2.42; 95%CI, 1.31–4.50; P=0.005) had a significantly higher odds ratio of SPD (**Table 3**). We found that economic status (e.g., income level, employment type) or social isolation (e.g., unmarried) were not associated with a higher prevalence of SPD among this population.

**Table 3.**
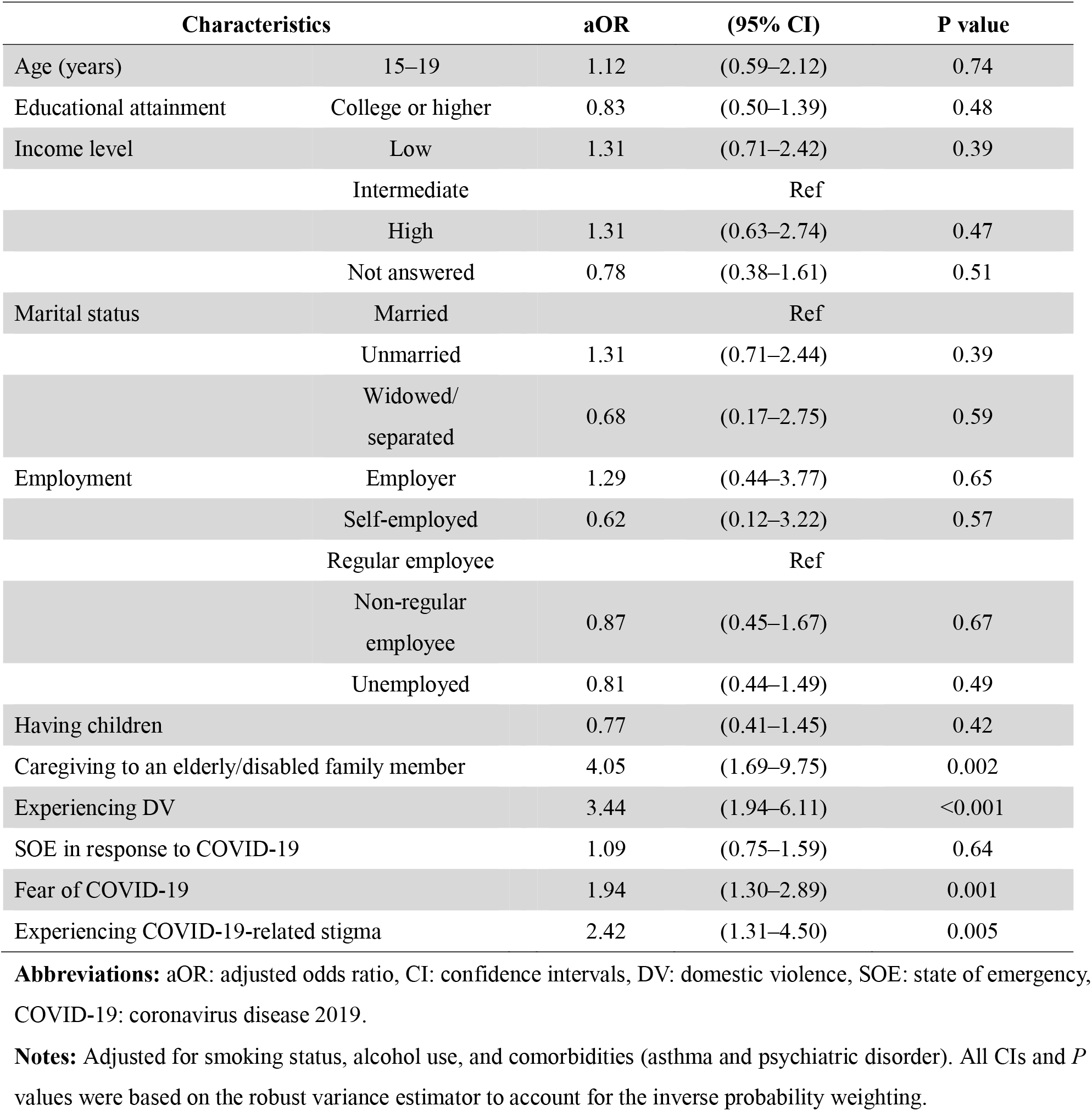
Factors associated with serious psychological distress among those aged 15–29 years (population with the highest risk of suicide during the COVID-19 pandemic)

## DISCUSSION

Using data from a nationally representative survey conducted during the COVID-19 pandemic, we identified several important factors that were associated with a higher rate of SPD in Japan. We found that women, younger age, income level, employment type, caregiving status, DV experience, and fear of COVID-19 were independently associated with SPD. Among young women—who are exhibiting an increased suicide rate during the COVID-19 pandemic in Japan—experiencing DV, fear of COVID-19, and COVID-19-related stigma were risk factors of SPD. On the other hand, economic situation and social isolation—factors considered to be causes of an increased suicide rate among this population—were not associated with the rate of SPD. Taken together, these findings indicate that underlying reasons for SPD may vary based on subgroups of the population, and therefore, targeted interventions based on age and gender may be more effective in mitigating negative impacts of the COVID-19 pandemic on the population’s mental health.

There are several potential mechanisms that may explain the factors associated with SPD in Japan. First, economic downturns due to the COVID-19 pandemic may negatively impact the mental health condition of the population. For example, we found that having a low-income level and being self-employed were associated with SPD, suggesting that the individuals in the general public were potentially vulnerable population affected by economic stagnation itself and social distancing due to the COVID-19 pandemic.^35,36^ Second, social isolation due to the SOE may cause a heavier burden of caregiving as well as DV from partners, resulting in mental illness. In our study, caregiving to an elderly/disabled family member was associated with SPD in both the general public and young women. It is known that reported cases of DV increased while people were staying home,^37^ probably leading to worse mental health conditions. Finally, as COVID-19 spread and society dramatically changed, fears about COVID-19 gradually developed making people psychologically unstable. Our findings, in which fear of COVID-19 was associated with SPD in all analyses, support this hypothesis.

Evidence is limited as to which factors affected mental health status at the population level during the COVID-19 pandemic. Wang et al. conducted a systematic review and meta-analysis examining the factors associated with psychological distress (i.e., anxiety and depression) in the general population, and found that women, younger age, and lower SES were associated with both anxiety and depression.^10^ Regarding gender- and age-related risk factors, Rehm and Shield reported that anxiety and depression are more prevalent in women than in men.^13^ In addition to such pre-existing psychological distress, previous studies reported that women in preconception,^38^ pregnancy,^39^ or postpartum periods^40^ experienced anxiety or depressive symptoms during the COVID-19 pandemic. While informative, evidence regarding young women in the factors associated with SPD is limited. To our knowledge, this is the first study that has examined the risk factors of SPD comprehensively in both the general public and the mentally vulnerable population (young women) using large-scale, nationally representative data.

Our study has limitations. First, because this survey was conducted between August and September 2020, it is possible that our findings would have been different if more recent data had been used. However, the data was collected just before October, 2020, when suicides among young women actually increased in Japan,^41^ and therefore, our findings may reflect the most critical period regarding the underlying reasons for increased suicide rates in Japan. Second, due to the self-reported design of our study, not all the variables were based on the validated questionnaire. For example, COVID-19-related stigma, which was an important risk factor for SPD among young women, was collected via a single-item questionnaire we developed, resulting in a lack of detailed stigma profiles.^42^ However, our survey used the items (K-6 scale), the reliability and validity of which have been tested and verified in prior studies.^31,43^ Third, given that our data were collected using an internet-based survey, respondents may be different in meaningful ways than those who were not recruited. However, we used propensity-score weighted analysis to adjust for the demographic, socio-economic, and health-related differences between respondents of the present study and the Japanese general public.^33,34^ Finally, our findings may not be generalizable to populations in other countries.

Using large-scale, nationally representative survey data, we identified several important factors associated with SPD during the COVID-19 pandemic in Japan’s general population. In particular, we found that female gender, younger age, income level, employment type, caregiving status, DV experience, and fear of COVID-19 were key risk factors for a higher rate of SPD. Among young women who presented higher suicide rates during the COVID-19 pandemic in Japan, experience of DV, fear of COVID-19, and COVID-19-related stigma were risk factors associated with SPD. Our findings highlight the importance of designing countermeasures based on the age and gender of the population to more effectively mitigate negative impacts on mental health during the COVID-19 pandemic.

## Supporting information

Supplementary content

## Data Availability

The data used in this study are not available in a public repository because they contain personally identifiable or potentially sensitive participants' information. Based on the regulations for ethical guidelines in Japan, the Research Ethics Committee of the Osaka International Cancer Institute has imposed restrictions on the dissemination of the data collected in this study.

## Acknowledgments

The JACSIS study was supported by the Japan Society for the Promotion of Science (JSPS) KAKENHI Grants [grant number 17H03589;19K10671;19K10446;18H03107; 18H03062; 19H03860], the JSPS Grant-in-Aid for Young Scientists [grant number 19K19439], Research Support Program to Apply the Wisdom of the University to tackle COVID-19 Related Emergency Problems, University of Tsukuba, and Health Labour Sciences Research Grant [grant number 19FA1005;19FG2001; 19FA1012] and the Japan Agency for Medical Research and Development (AMED; grant number 2033648). We would like to appreciate Drs. Kota Katanoda, Keisuke Kuwahara, Kanami Tsuno, Kenji Takeuchi, Hiroshi Murayama, Ai Hori, Isao Muraki, Naoki Kondo, and Takeo Fujiwara for the support related to the data collection.

## Author contributions

Dr. Yoshioka had full access to all the data in this study and takes responsibility for the integrity of the data and the accuracy of the data analysis.

### Concept and design

Yoshioka, Okubo, Tabuchi, and Tsugawa. *Acquisition, analysis, or interpretation of the data:* All authors. *Drafting of the manuscript:* Yoshioka, Tsugawa.

### Critical revision of the manuscript for important intellectual content

All authors.

### Statistical analysis

Yoshioka.

### Administrative, technical, or material support

Okubo, Tabuchi, Odani, Shinozaki, Tsugawa.

### Supervision

Tsugawa.

## SUPPLEMENT

### Supplementary Method

**Supplementary Table 1. Baseline characteristics of unweighted respondents**

**Supplementary Table 2. Characteristics of respondents among those aged 15-29 years (population with the highest risk of suicide during the COVID-19 pandemic)**

