## Supplementary content for "Factors Associated with Serious Psychological Distress during the COVID-19 Pandemic in Japan"

**Supplementary Method**

**Supplementary Table 1. Baseline characteristics of unweighted respondents**

**Supplementary Table 2. Characteristics of respondents among those aged 15-29 years (population with the highest risk of suicide during the COVID-19 pandemic)**

**Supplementary Method**

***Declaration of state of emergency in Japan***

The Japanese government had declared a state of emergency (SOE) from 7 April to 25 May 2020. The SOE authorizes the prefectural governors to request residents of a given prefecture to refrain from leaving their homes for non-essential reasons. All 47 prefectures in Japan were subject to the SOE during this period. Also, 13 prefectures (Hokkaido, Ibaraki, Saitama, Chiba, Tokyo, Kanagawa, Ishikawa, Gifu, Aichi, Kyoto, Osaka, Hyogo, and Fukuoka) were designated as the special alert prefectures. The residents of the special alert prefectures were forced to be confined to home longer than those in the other prefectures. In this study, we defined respondents influenced by the SOE in response to COVID-19 as the residents in these 13 prefectures.

***Definition of domestic violence (DV) and COVID-19-related stigma***

The survey included specific questions developed for identifying domestic violence and COVID-19-related stigma.

The survey items used to identify DV were:

- “From April 2020 to the present, have you been physically assaulted, such as punched, kicked, had objects thrown at you, or been locked in a room?”
- “From April 2020 to the present, have you been subjected to behaviors that hurt your self-esteem, such as being verbally abused, being told unpleasant things, or being ignored for a long time?”
- “From April 2020 to the present, have you ever had your savings or pension taken away from you without your consent?”
- “From April 2020 to the present, have you ever experienced non-consensual sex?”
- “From April 2020 to the present, have you ever had anxiety about an unwanted pregnancy?”

The response options were “Yes” and “No.” If any one of the above questions was answered “Yes,” we defined the respondents as experiencing DV.

The COVID-19-related stigma was defined using the following item included in the survey:

- “From April 2020 to the present, have you felt discriminated against or prejudiced related to COVID-19?”

The response options were “Yes” and “No.” If the respondents answered “Yes,” we defined them as experiencing COVID-19-related stigma.

**Supplementary Table 1. Baseline characteristics of unweighted respondents**

| **Characteristics** | |  | **Overall**  **n = 25,482** | |
| --- | --- | --- | --- | --- |
| Women |  |  | 12,845 | (50.4) |
| Age (years) | 15–29 |  | 4,425 | (17.4) |
|  | 30–44 |  | 6,071 | (23.8) |
|  | 45–59 |  | 6,846 | (26.9) |
|  | 60–79 |  | 8,140 | (31.9) |
| Educational attainment | High-school-educated or lower |  | 8,353 | (32.8) |
|  | College-educated or higher |  | 17,129 | (67.2) |
| Income level | Low |  | 6,820 | (26.8) |
|  | Intermediate |  | 6,902 | (27.1) |
|  | High |  | 6,486 | (25.5) |
|  | Unknown/declined to answer |  | 5,274 | (20.7) |
| Marital status | Married |  | 15,230 | (59.8) |
|  | Unmarried |  | 7,806 | (30.6) |
|  | Widowed/ separated |  | 2,446 | (9.6) |
| Employment | Employer |  | 847 | (3.3) |
|  | Self-employed |  | 1,645 | (6.5) |
|  | Regular employee |  | 8,666 | (34.0) |
|  | Non-regular employee |  | 4,296 | (16.9) |
|  | Unemployed |  | 10,028 | (39.4) |
| Having children | |  | 8,583 | (33.7) |
| Caregiving to an elderly/disabled family member | |  | 1,923 | (7.5) |
| Experiencing DV | |  | 1,976 | (7.8) |
| SOE in response to COVID-19 | |  | 15,925 | (62.5) |
| Fear of COVID-19 | |  | 11,488 | (45.1) |
| Experiencing COVID-19-related stigma | |  | 867 | (3.4) |
| Smoking status | Never |  | 14,225 | (55.8) |
|  | Ever |  | 7,854 | (30.8) |
|  | Current |  | 3,403 | (13.4) |
| Alcohol use | Never |  | 5,537 | (21.7) |
|  | Ever |  | 7,788 | (30.6) |
|  | Current |  | 12,157 | (47.7) |
| Comorbidities | Hypertension |  | 4,599 | (18.0) |
|  | Diabetes |  | 1,565 | (6.1) |
|  | Asthma/COPD |  | 896 | (3.5) |
|  | Cardiovascular disease |  | 509 | (2.0) |
|  | Stroke |  | 245 | (1.0) |
|  | Cancer |  | 455 | (1.8) |
|  | Psychiatric disorders |  | 1,449 | (5.7) |

**Abbreviations.** CI: confidence intervals, COPD: chronic obstructive pulmonary disease, DV: domestic violence, SOE: state of emergency, COVID–19: coronavirus disease 2019.

**Notes.** All proportions are expressed as numbers and percentages (%).

**Supplementary Table 2. Characteristics of respondents among those aged 15-29 years (population with the highest risk of suicide during the COVID-19 pandemic)**

| **Characteristics** |  |  | **Unweighted**  **n = 2,295** | | **Weighted**  **n = 2,295** | |
| --- | --- | --- | --- | --- | --- | --- |
| Age (years) | 15–19 |  | 715 | (31.2) | 715 | (31.2) |
|  | 20–29 |  | 1580 | (68.8) | 1580 | (68.9) |
| Academic attainment | High-school-educated or lower |  | 730 | (31.8) | 1380 | (60.1) |
|  | College-educated or higher |  | 1565 | (68.2) | 915 | (39.9) |
| Income level | Low |  | 636 | (27.7) | 626 | (27.3) |
|  | Intermediate |  | 544 | (23.7) | 487 | (21.2) |
|  | High |  | 438 | (19.1) | 351 | (15.3) |
|  | Unknown/declined to answer |  | 677 | (29.5) | 831 | (36.2) |
| Marital status | Married |  | 421 | (18.3) | 452 | (19.7) |
|  | Unmarried |  | 1849 | (80.6) | 1794 | (78.2) |
|  | Widowed/separated |  | 25 | (1.1) | 49 | (2.1) |
| Employment | Employer |  | 53 | (2.3) | 45 | (2.0) |
|  | Self-employed |  | 24 | (1.0) | 20 | (0.9) |
|  | Regular employee |  | 717 | (31.2) | 648 | (28.2) |
|  | Non-regular employee |  | 318 | (13.9) | 454 | (19.8) |
|  | Unemployed |  | 1183 | (51.5) | 1128 | (49.2) |
| Having children |  |  | 296 | (12.9) | 403 | (17.6) |
| Caregiving to an elderly/disabled family member | |  | 84 | (3.7) | 84 | (3.7) |
| Experiencing DV | |  | 265 | (11.5) | 307 | (13.4) |
| SOE in response to COVID-19 | |  | 1517 | (66.1) | 897 | (39.1) |
| Fear of COVID-19 | |  | 998 | (43.5) | 1086 | (47.3) |
| Experiencing COVID-19-related stigma | |  | 146 | (6.4) | 102 | (4.4) |
| Smoking status | Never |  | 1937 | (84.4) | 1855 | (80.8) |
|  | Ever |  | 285 | (12.4) | 333 | (14.5) |
|  | Current |  | 73 | (3.2) | 107 | (4.7) |
| Alcohol use | Never |  | 926 | (40.3) | 1000 | (43.6) |
|  | Ever |  | 786 | (34.2) | 743 | (32.4) |
|  | Current |  | 583 | (25.4) | 552 | (24.1) |
| Comorbidities | Asthma |  | 88 | (3.8) | 123 | (5.4) |
|  | Psychiatric disorders |  | 173 | (7.5) | 179 | (7.8) |

**Abbreviations.** CI: confidence intervals, COPD: chronic obstructive pulmonary disease, DV: domestic violence, SOE: state of emergency, COVID–19: coronavirus disease 2019.

**Notes.** All proportions are expressed as numbers and percentages (%). In the weighted group, the sum of respondents did not necessarily equal the number of categories because of sample weighting and rounding. The weights were predicted from the logistic model including area (residence), marital status, education, home-ownership (household), self-rated health, and smoking status were adjusted for in individuals aged 20-29 years; and area, education, home-ownership (household) and self-rated health were adjusted for (omitting marital and smoking status) in individuals aged 15-19 years.
